# Attendance-related healthcare resource utilisation and costs in patients with long QT syndrome in Hong Kong: A retrospective cohort study

**DOI:** 10.1101/2022.11.12.22282256

**Authors:** Cheuk To Chung, Oscar Hou In Chou, Teddy Tai Loy Lee, Danny Radford, Kamalan Jeevaratnam, Wing Tak Wong, Shuk Han Cheng, Ngai Shing Mok, Tong Liu, Gary Tse, Sharen Lee

## Abstract

**Introduction:** The understanding of healthcare resource utilisation and its related costs is crucial for optimizing resource allocation in the healthcare setting. There is currently a paucity of published studies investigating healthcare costs related to long QT syndrome (LQTS).

**Method:** This was a retrospective study of LQTS patients from Hong Kong, China. The healthcare resource utilisation for Accident and Emergency (A&E), inpatient and specialist outpatient settings across a 19-year period was extracted and analysed. Costs in US dollars were calculated using unit costs.

**Results:** The cohort consists of 125 LQTS patients with a mean presentation age of 26.7 ± 22.0 years old. Of these, 45 patients presented with ventricular tachycardia/ventricular fibrillation (VT/VF) and 44 patients had an implantable cardioverter-defibrillator (ICD) implementation. At the individual patient level, the median annualised costs were $69 (30-183) at the A&E setting, $10270 (2248-64006) at the inpatient setting and $675 (393-1329) at the special outpatient setting. Patients who presented with VT/VF initially had significantly higher annualised median costs in the inpatient ($59843 [13812-214930] vs. $5480 [1162-23111], p<0.0001) and specialist outpatient setting ($823 [539-1694] vs $609 [383-1269], p=0.133) compared to patients without VT/VF initially.

**Conclusion:** There is an increasing healthcare demand in the inpatient and specialist outpatient settings for LQTS patients. The most expensive attendance type was inpatient setting stay at $10270 per year. The total median annualised cost of LQTS patients without VT/VF was 90% lower compared to patients with VT/VF.

## Introduction

Long QT syndrome (LQTS) is a type of cardiac ion channelopathy that is characterised by the prolongation of QT interval > 480ms and T-wave abnormalities displayed on the electrocardiogram (ECG). Congenital LQTS can be caused by mutations in genes encoding for cardiac ion channels responsible for mediating action potential conduction or repolarisation ^1^. The three types of LQTS are typically associated with the mutation of three main causative genes encoding for ion channels: *KCNQ1* encoding for potassium channel proteins Kv7.1 in LQTS type 1 (LQT1); *KCNH2* encoding for potassium channel proteins Kv11.1 in LQTS type 2 (LQT2); and *SCN5A* encoding for sodium channels Nav.1.5 in LQTS type 3 (LQT3) ^2^. Common symptoms of LQTS include syncope, palpitation and seizure. Current research suggests that LQTS is a leading cause of sudden cardiac death (SCD) ^3^, though the risk of SCD varies between individuals depending on gender, age and genotype ^4^. In addition, the condition can be impacted by the patient’s drug usage and comorbidities such as diabetes mellitus ^5, 6^. Currently, there is no definitive cure for LQTS, however the use of implantable cardioverter defibrillators, beta blockers coupled with lifestyle modifications may alleviate symptoms and reduce the risk of adverse cardiac events ^1^.

There is currently an abundance of publications examining the genetic basis of LQTS. Amongst Asian populations, while some studies have postulated the genotype-phenotype correlation of LQTS, there are fewer to no studies analyzing its economic burden. This may be attributed to the lack of adequate data and clinical trials for analysis. The Gly387Arg variant of the *KCNJ5* gene has shown an association with congenital LQTS (cLQTS) in a multinational Chinese pedigree ^7^. Another retrospective study from Hong Kong consisting of 121 cLQTS patients identified novel mutations in 7 putative ion channel genes ^8^. To corroborate, another study found that the majority of LQTS patients from a Thai population were symptomatic, in which LQT2 and LQT3 patients developed symptoms during sleep or at rest ^9^.

Although the genetic basis of LQTS has been extensively studied, the literature surrounding the economic burden and clinical management of this disease remains scarce. Several studies have examined the cost-effectiveness of early genetic testing and neonatal ECG screening in western populations ^10-12^. Moreover, healthcare resource utilisation and related costs may differ within the LQTS population, as affected patients range in varying levels of disease severity. Hence, there is increasing demand for services such as diagnostic testing and subsequent therapeutic approaches. With a better understanding of resource allocation and healthcare expenditure, this may facilitate the provision of more timely treatment for patients and better healthcare policies. Thus, this warrants the importance of an investigation of the healthcare resource utilisation of LQTS patients. In this study, we investigated the attendance-related healthcare resource utilisation and its costs for LQTS patients from Hong Kong, China.

## Methods

### Study population

This study on cardiac arrhythmias was approved by The Joint Chinese University of Hong Kong-New Territories East Cluster Clinical Research Ethics Committee. The cohort included consecutive patients diagnosed with LQTS between January 1^st^, 1997 to December 31^st^, 2019 in public hospitals or clinics under the Hong Kong Hospital Authority. This system has been used previously by our team to investigate rare congenital arrhythmic syndromes ^13-15^ as well as common diseases ^16-18^. The patient identification and data extraction process involved reviewing centralised electronic health records from public hospitals. Studies on risk prediction using this cohort have already been published ^8, 19^. This study focuses on costs of attendances but also illustrate the baseline clinical characteristics for information purposes. While the initial diagnosis was made by case physicians, they were later verified by G.T. through the inspection of documented ECGs, genetic reports, case notes and diagnostic test results.

### Clinical and electrocardiographic data collection

The baseline clinical data extracted from the electronic health records in-cooperates: 1) sex; 2) age of first characteristic ECG presentation and last follow-up; 3) follow-up duration; 4) family history of SCD and the specific ion channelopathy; 5) syncope manifestation and its frequency; 6) presentation of sustained VT/VF and its frequency; 7) performance of electrophysiological study (EPS), 24-hours Holter study, ion channelopathy-specific genetic testing, and the respective results; 8) performance of echocardiogram; 9) presence of other arrhythmias; 9) implantation of implantable cardioverter-defibrillator (ICD); 10) occurrence, cause and age of death; 11) period between the initial presentation of characteristic ECG and the first post-diagnosis ventricular tachycardia/ ventricular fibrillation (VT/VF) episode; 12) initial disease manifestation (asymptomatic, syncope, VT/VF).

In this study, the phenotypic presentation of LQTS can be defined as the presence of VT/VF, syncope or a history of cardiac events. Spontaneous VT/VF refers to VT/VF during follow up, and incidental VT/VF refers to VT/VF episodes that were not induced iatrogenically. Asymptomatic patients denote the absence of all symptoms. Other conditions and symptoms including atrioventricular block, atrial tachyarrhythmias, and palpitations were also taken into consideration. The presence of positive EPS is defined as the induction of VT/VF that either sustained a minimum of 30 seconds or produced hemodynamic collapse. The baseline ECG was extracted and documented at the earliest time possible after the presentation of an initial characteristic ECG pattern.

### Statistical analysis

The Fisher’s exact test was used to compare categorical variables, expressed as a total number and percentage, while the t-test was used to compare continuous variables and expressed as mean± standard deviation. Statistical significance was defined as a P-value of less than 0.05. The Mann-Whitney-Wilcoxon Test was also performed to validate the statistical significance of subgroup analysis. All statistical analysis was performed using R Studio (Version: 1.3.1073).

### Healthcare utilisation and cost analyses

Healthcare resource utilisation for accident and emergency (A&E), inpatient, general outpatient and specialist outpatient attendances were analysed over a 23-year period (1997-2019). The costs for these attendances were calculated using unit costs published by the local government and then later annnualised. The cost values are presented in US Dollars. A further comparison was conducted by stratifying the cohort into different subgroups based on three variables: the initial presentation of VT/VF, syncope and the conduction of a genetic test. In addition, the incidence rate ratio (IRR) was also calculated to compare two subgroups: (1) patients with and without syncope; (2) patients who presented with syncope initially with and without incidental VT/VF. The IRR for the former subgroup was calculated by dividing the patients with syncope total person per year by patients with no syncope total person per year. The latter was performed as a form of sensitivity analysis to take into consideration the potential differences in their risk profiles as a result of VT/VF.

## Results

### Baseline clinical characteristics, ECG features and arrhythmic features

In this retrospective study, 125 patients with LQTS were included with a mean follow-up duration of 96 months. The mean presentation age of the cohort was 26.7 years old and 73 patients were female. During the follow up period, both patients with and without initial VT/VF subgroups had 16 patients exhibit incident VT/VF during follow up. Patients without initial VT/VF had a greater occurrence of syncope compared to patients with initial VT/VF (48 vs. 20). In regard to family background, 49 patients presented a family history of LQTS and 19 patients with a family history of VT/VF or SCD. In contrast, only 4 patients performed an EPS. In regards to baseline ECG characteristics, patients with an initial presentation of VT/VF demonstrated a significantly lower max QTc interval during recovery (275.0ms vs 516.3ms) and greater T axis (61.0° vs 51.2°) compared to their counterparts. The baseline characteristics of the whole LQTS cohort, including a comparison of LQTS patients with and without an initial presentation of VT/VF, are summarised in **Table 1**.

**Table 1.** Baseline characteristics of the study cohort. Categorical and continuous variables were compared between LQTS patients with and without an initial presentation of VT/VF using Fisher’s exact test and t-test, respectively. Bolded text indicates P<0.05.

| Variable | All LQTS patients (n=125) | LQTS patients with initial VT/VF (n=29) | LQTS patients without initial VT/VF (n=96) | p-value |
| --- | --- | --- | --- | --- |
| **Clinical characteristics** |  |  |  |  |
| Female | 73 (58.4) | 16 (55.2) | 57 (59.4) | 0.830 |
| Presentation age (years) | 26.7±22.0 | 32.7±22.2 | 25.0±26.0 | **<0.0001** |
| Follow up duration (months) | 96.0±64.6 | 111.7±76.2 | 91.2±60.2 | **<0.0001** |
| Schwartz score criteria | 4.3±1.2 | 4.5±0.9 | 4.2±1.2 | **<0.0001** |
| Family history of LQTS | 49 (39.2) | 4 (13.8) | 45 (46.9) | **<0.01** |
| Family history of VT/VF/SCD | 19 (15.2) | 2 (6.9) | 17 (17.7) | 0.238 |
| Initial syncope | 64 (51.2) | 18 (62.1) | 46 (47.9) | 0.208 |
| Syncope | 68 (54.4) | 20 (69.0) | 48 (50) | 0.090 |
| No. of syncope episodes | 1.1±1.5 | 1.2±1.3 | 1.1±1.5 | 0.386 |
| Palpitations | 19 (15.2) | 3 (10.3) | 16 (16.7) | 0.559 |
| Initial VT/VF | 29 (23.2) | 29 (100) | 0 (0) | **<0.0001** |
| Spontaneous VT/VF during follow up | 45 (36) | 28 (96.6) | 17 (17.7) | **<0.0001** |
| No. of spontaneous VT/VF episodes | 2.5±13.9 | 2.3±2.4 | 2.6±15.8 | **0.005** |
| High VT/VF burden (≥2 episodes) | 26 (20.8) | 14 (48.3) | 12 (12.5) | **<0.001** |
| Incident VT/VF | 32 (25.6) | 16 (55.2) | 16 (16.7) | **<0.001** |
| No. of SVE/SVT/AT/AF/sick sinus/AVB episodes | 0.2±0.8 | 0.5±1.5 | 0.1±0.4 | 0.105 |
| Performance of treadmill test | 45 (36) | 4 (13.8) | 41 (42.7) | **0.004** |
| Exercise-recovery-induced QT prolongation in treadmill | 31 (24.8) | 0 (0) | 31 (32.3) | **<0.001** |
| Genetic test | 77 (61.6) | 13 (44.8) | 64 (66.7) | **0.049** |
| EPS | 4 (3.2) | 1 (3.4) | 3 (3.1) | 1 |
| *Induced VT/VF under EPS* | 2 (1.6) | 0 (0) | 2 (2.1) | 1.00 |
| ICD | 44 (35.2) | 25 (86.2) | 19 (19.8) | **<0.0001** |
| Number of Holter conducted | 0.5±0.9 | 0.8±1.4 | 0.4±0.6 | 0.441 |
| **Baseline ECG Characteristics** |  |  |  |  |
| PVC | 23 (18.4) | 10 (34.5) | 13 (13.5) | **0.026** |
| TdP | 33 (26.4) | 21 (72.4) | 12 (12.5) | **<0.0001** |
| Initial ECG QTc (ms) | 502.9±44.0 | 511.8±39.1 | 496.2±45.2 | **<0.0001** |
| Heart rate (bpm) | 77.2±23.6 | 71.8±16.7 | 79.1±25.5 | **<0.0001** |
| P-wave duration (ms) | 103.9±16.0 | 112.3±19.3 | 100.5±13.6 | **<0.0001** |
| PR interval (ms) | 160.8±30.7 | 163.7±19.3 | 159.8±31.0 | **<0.0001** |
| QRS interval (ms) | 97.3±23.2 | 99.6±26.5 | 96.4±22.0 | **<0.0001** |
| QT interval (ms) | 442.6±71.3 | 454.1±61.7 | 438.2±74.6 | **<0.0001** |
| QTc Interval (ms) | 488.4±45.2 | 497.2±41.7 | 485.6±46.2 | **<0.0001** |
| Max QTc interval during recovery (ms) | 502.9±112.2 | 275.0±388.9 | 516.3±73.8 | 0.501 |
| P axis | 54.9±42.7 | 63.2±48.3 | 51.7±40.4 | **<0.0001** |
| QRS axis | 56.1±59.8 | 67.0±84.4 | 52.0±47.4 | **<0.0001** |
| T axis | 53.9±56.9 | 61.0±76.8 | 51.2±47.7 | **<0.0001** |
| R wave in lead V5 | 1.1±0.7 | 1.1±0.6 | 1.2±0.7 | 0.728 |
| S wave in lead V1 | 0.7±0.4 | 0.8±0.4 | 0.6±0.4 | 0.078 |

LQTS: Long QT syndrome; SCD: sudden cardiac death; TdP: Torsades de pointes; PVC: premature ventricular contractions; VT/VF: ventricular tachycardia/fibrillation; EPS: electrophysiological study; QTc: corrected QT interval; NSVT: non-sustained ventricular tachycardia; ICD: implantable cardioverter-defibrillator; ECG: electrocardiogram; SVE: sinus with Supraventricular Ectopy; SVT: supraventricular tachycardia; AT: atrial tachycardia; AF: atrial fibrillation; AVB; atrioventricular block

### Healthcare resource utilisation and costs

The total number of attendances for A&E, inpatient and specialist outpatient settings in the cohort were 1130, 1253 and 9121 respectively, corresponding to total costs of $178,650, $182,267,011 and $1,395,114. At the single-patient level, the median (IQR) number of attendances for A&E, inpatient and specialist outpatient settings were 5 (2-11), 5 (2-10) and 40 (21-89), respectively. The corresponding total median cost was $790 (316-1739) for A&E, $101,579 (23,599-590,953) for inpatient and $6118 (3,212-13613) for specialist outpatient. Furthermore, the median annualised costs for each setting are as follows: $69 (30-183) for A&E, $10,270 (2,248-64,006) for inpatient and $675 (393-1,329) for specialist outpatient. A summary of the cost analysis of the whole cohort is illustrated in **Table 2**.

**Table 2.** All cause LQTS healthcare utilisation and costs. Median (25th to 75th percentile) values are presented. Costs are shown in US dollars.

| **Attendance type** | **Attendances** | **Costs (\$)** | **Annualised costs (\$/year)** |
| --- | --- | --- | --- |
| Accident & Emergency | 5 (2-11) | 790 (316-1739) | 69 (30-183) |
| Inpatient | 5 (2-10) | 101579 (23599-590953) | 10270 (2248-64006) |
| Specialist outpatient | 40 (21-89) | 6118 (3212-13613) | 675 (393-1329) |

The attendance costs comparison of the three sets of subgroups is summarised in **Table 3-5**. The overall median attendance for patients without iniital VT/VF and genetic test performed was relatively lower in inpatient and specialist outpatient settings compared to their respective counterparts. Contrarily, patients with syncope had overall higher attendance in all settings compared to patients without syncope. The annualised median costs for patients who presented with VT/VF is significantly higher in the inpatient ($59843 vs $5480) and specialist outpatient setting ($823 vs $609). Following on, patients who did not undergo genetic tests indicated overall greater annualised median costs in A&E ($81 vs $69), inpatient ($27,435 vs $5356) and specialist outpatient setting ($928 vs $560). Furthermore, patients with syncope show a greater annualised median cost in A&E ($76 vs $62) and inpatient setting ($10,275 vs $9246). However, patients without syncope show a greater annualised cost in the specialist outpatient setting ($834 vs $536).

**Table 3.** Comparison of healthcare utilisation and costs between LQTS patients with and without an initial presentation of VT/VF. Median (25th to 75th percentile) values are presented. Costs are shown in US dollars.

| Attendance type | Attendance | | | Costs (\$) | | | Annualised costs (\$/year) | | |
| --- | --- | --- | --- | --- | --- | --- | --- | --- | --- |
|  | Initial VT/VF | No initial VT/VF | *p-value* | Initial VT/VF | No initial VT/VF | *p-value* | Initial VT/VF | No initial VT/VF | *p-value* |
| **Accident & Emergency** | 5 (2-11) | 5 (2-11) | 0.948 | 790<br>(316-1739) | 790<br>(316-1739) | 0.948 | 84 (30-171) | 68 (30-189) | 0.667 |
| **Inpatient** | 6 (3-11), | 4 (2-9) | 0.232 | 541766<br>(179915-1031116) | 61128<br>(12981-255574) | **<0.0001** | 59843<br>(13812-214930) | 5480<br>(1162-23111) | **<0.0001** |
| **Specialist outpatient** | 45 (7-103) | 37 (20-81) | 0.216 | 6883<br>(4130-15754) | 5659<br>(3059-12428) | 0.216 | 823<br>(539-1694) | 609 (383-1269) | 0.133 |
| **All** | 56 (32-125) | 46 (24-101) | 0.306 | 549439<br>(184361-1048610) | 67578<br>(16356-269740) | **<0.0001** | 60750<br>(14381-216795) | 6158<br>(1575-24569) | **<0.0001** |

**Table 4.** Comparison of healthcare utilisation and costs between patients who performed genetic test and those who did not. Median (25th to 75th percentile) values are presented. Costs are shown in US dollars.

| Attendance type | Attendance | | | Costs (\$) | | | Annualised costs (\$/year) | | |
| --- | --- | --- | --- | --- | --- | --- | --- | --- | --- |
|  | Genetic test performed | No genetic test performed | *p-value* | Genetic test performed | No genetic test performed | *p-value* | Genetic test performed | No genetic test performed | *p-value* |
| **Accident & Emergency** | 4 (2-8) | 7 (3-13) | 0.178 | 632 (316-1265) | 1028 (435-2055) | 0.178 | 69 (27-184) | 81 (38-201) | 0.777 |
| **Inpatient** | 4 (1-7) | 7 (4-15) | **<0.01** | 53262<br>(11362-205234) | 326725<br>(103710-1508566) | **<0.0001** | 5356<br>(1208-29162) | 27435<br>(6192-214392) | **<0.01** |
| **Specialist outpatient** | 32 (19-52) | 74 (33-130) | **<0.001** | 4895<br>(2906-7954) | 11319<br>(4971-19923) | **<0.001** | 560 (342-914) | 928 (568-1628) | **<0.01** |
| **Total** | 40 (22-67) | 88 (39-158) | **<0.01** | 58789<br>(14585-214453) | 339072<br>(109116-1530543) | **<0.0001** | 5985<br>(1577-30260) | 28444<br>(6798-216221) | **<0.001** |

**Table 5.** Comparison of healthcare utilisation and costs between patients with and without syncope. Median (25th to 75th percentile) values are presented. Costs are shown in US dollars.

| Attendance type | Attendance | | | Costs (\$) | | | Annualised costs (\$/year) | | |
| --- | --- | --- | --- | --- | --- | --- | --- | --- | --- |
|  | Syncope | No Syncope | *p-value* | Syncope | No Syncope | *p-value* | Syncope | No Syncope | *p-value* |
| **Accident & Emergency** | 6 (2-13) | 4 (2-8) | 0.188 | 949 (316-2095) | 632 (316-1265) | 0.188 | 76 (42-202) | 62 (26-143) | 0.149 |
| **Inpatient** | 5 (3-10) | 4 (1-10) | 0.313 | 101757 (40527-591415) | 90900 (5353-445758) | 0.358 | 10275 (3091-60035) | 9246 (367-57015) | 0.369 |
| **Specialist outpatient** | 41 (22-83) | 38 (19-89) | 0.905 | 6195 (3327-12657) | 5812 (2906-13613) | 0.905 | 563 (364-965) | 834 (493-1665) | 0.080 |
| **All** | 52 (27-106) | 46 (22-107) | 0.772 | 108900 (44170-606167) | 97345 (8576-460636) | 0.414 | 10914 (3497-61202) | 10142 (886-58823) | 0.955 |

The IRR and the respective confidence intervals (CIs) for patients with and without syncope were also calculated, which is detailed in **Table 6**. Patients with syncope were associated with increased costs compared to patients without syncope in the A&E (IRR: 6.13 [2.95-12.72]) and specialist outpatient setting (IRR: 1.77 [1.47-2.14]). In addition, patients who presented with syncope and incidental VT/VF were associated with lower costs compared to patients who presented with syncope initially but without incidental VT/VF **(Table 7)**.

**Table 6.**
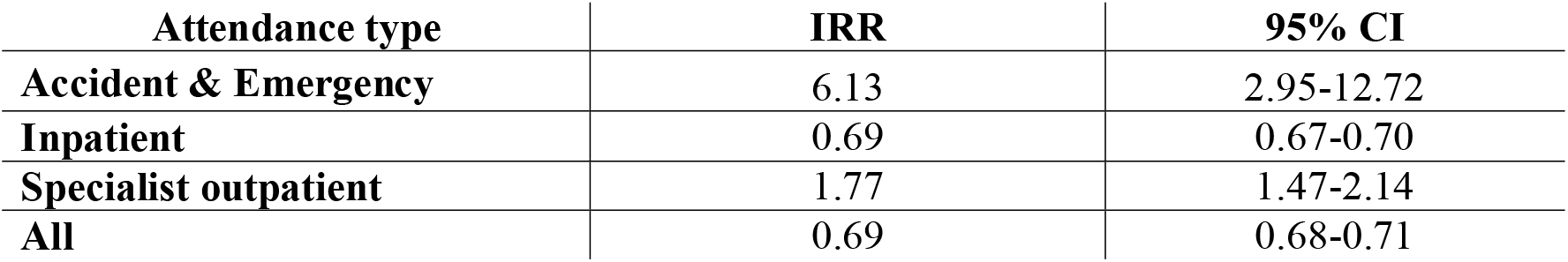
Incidence rate ratios of annualised costs for patients with and without syncope

**Table 7.** Sensitivity analysis comparing annualised costs of patients who presented with syncope initially with and without incidental VT/VF

| Attendance type | IRR | 95% CI |
| --- | --- | --- |
| **Accident & Emergency** | 0.13 | 0.07-0.25 |
| **Inpatient** | 0.37 | 0.36-0.38 |
| **Specialist outpatient** | 0.10 | 0.07-0.13 |
| **All** | 0.36 | 0.35-0.37 |

## Discussion

This is the first territory-wide cohort study examining healthcare resource utilisation and its related costs for LQTS. Several major findings were identified in this study: (1) LQTS patients require more services from the specialist outpatient settings; (2) The most expensive attendance type was inpatient stay at $10270 per year, followed by outpatient setting at $675 and A&E at $69 per year; (3) The total median annualised costs of LQTS without an initial presentation VT/VF was 90% less compared to their counterparts. This notwithstanding, the present study also demonstrated important clinical characteristics that may correlate to increased risks of VT/VF amongst LQTS patients.

The present study demonstrates that the healthcare costs of LQTS are affected by the disease phenotype. Patients with syncope were associated with increased costs compared to patients without syncope in the A&E and specialist outpatient setting. This may be because patients with syncope are at greater risk for incident VT/VF and require an ICD implantation. However, amongst patients who presented with syncope initially, those with incidental VT/VF were associated with lower costs compared to patients without incidental VT/VF. The difference in costs between those with and without incidental VT/VF may be attributed to the increased frequency of follow up attendances and diagnostic tests. Coyle *et al*. found that catheter ablation was more cost-effective compared to escalated antiarrhythmic therapy for patients with amiodarone-refractory VT. Interestingly, there was no significant difference in the cost-effectiveness of the two interventions for patients with sotalol-refractory VT ^20^. Weiss *et al*. identified that healthcare expenditures among VT/VF patients who received a implanatable defribrillator to be consistently high due to greater demand for long-term inpatient care services, measuring a cost-effectiveness ratio of $78,400 per life-year gained ^21^. The additional costs of implantation and follow-up may explain the greater overall healthcare costs of LQTS patients with incidental VT in our study.

Current literature demonstrates firm evidence supporting the increased risks of VT/VF, SCD and syncope in LQTS patients. However, effective management and risk stratification of LQTS patients remains a difficult task ^22^. While the current guidelines recommend against an ICD implantation for patients with acquired LQTS ^23^, there has been preliminary evidence showing the success of appropriate shocks in 44% of patients ^24^. Patients and their at-risk family members often undergo an array of phenotypic and clinical assessments, in which genetic testing was concluded to be a moderately expensive approach ^11^. Philips *et al*. further extend these findings and found that an approximation of 2500 US dollars per year of life was saved using genetic testing compared to no testing of symptomatic index individuals, therefore postulating that early screening is cost-effective ^25^. On the other hand, Quaglini *et al*. compared the cost-effectiveness of routine mass ECG screening versus no screening strategy for 30,000 infants from an Italian population. The study found that neonatal ECG screening for LQTS is cost-effective and can prevent avoidable premature deaths ^12^. Among the observed studies of cost analyses, they were unable to account for the difference in frequencies and disease presentation amongst non-western population ethnicities ^26^, which could have a significant influence on treatment cost-effectiveness. The relevance of derived estimations toward Asian populations warrants further validation. Henceforward, the study’s findings may be applied to enhancing the efficacy of targeted screening and reduce costs. In the long term, optimised resource utilisation can reduce the patient’s financial burden and allow them to receive appropriate care.

### Strengths and limitations

This study presents several major strengths, this includes: (1) costs were estimated using unit costs across extended follow-up periods; (2) the use of a public, comprehensive electronic health record system from the city, combining attendances from multiple hospitals; (3) the inclusion of a large sample size can enhance the reliability of study findings.

However, some limitations should also be recognised. Firstly, due to the retrospective observational nature of the study, this suggests that the data interpretation may be susceptible to information and selection bias. However, given the majority of patients were closely followed-up via annual consultations, the bias is largely minimised by the detailed follow up and documentation. Secondly, the small sample size of the LQTS cohort limits the data reliability and extent of patient stratification as the condition is comparatively rare amongst other cardiac conditions in Hong Kong. However, it must be noted that our current cohort is already one of the largest cohorts among Asia. Moreover, as data regarding the family history of patients was not extensively detailed, this may raise some uncertainty about whether asymptomatic patients were family members of probands. Finally, cost analysis of the general outpatient setting was attempted but due to the low number of patients, any significant cost values could not be assessed. It is imperative to acknowledge that our cost analyses require additional validation in a prospective setting.

## Conclusion

To conclude, the present study demonstrated significant clinical characteristics and associated healthcare costs of LQTS patients from Hong Kong. These findings can offer novel insight into the refinement of healthcare interventions surrounding LQTS and other channelopathies. In the future, cost analysis could target smaller subgroups within the studied population.

## Data Availability

All data produced in the present study are available upon reasonable request to the authors

## Acknowledgements

None.

## Author Contributions

CTC —data acquisition, database interpretation, statistical analysis, cost analysis, manuscript drafting, manuscript revision

OHIC, TTLL, DR, KJ, WTW, SHC, NSM, TL, GT, SL – critical revision of manuscript, data analysis, data collection, manuscript drafting

## References

1. Shah SR, Park K and Alweis R. Long QT Syndrome: A Comprehensive Review of the Literature and Current Evidence. Curr Probl Cardiol. 2019;44:92–106.

2. Tester DJ and Ackerman MJ. Genetics of long QT syndrome. Methodist Debakey Cardiovasc J. 2014;10:29–33.

3. Steinberg C. Diagnosis and clinical management of long-QT syndrome. Curr Opin Cardiol. 2018;33:31–41.

4. Longo UG, Risi Ambrogioni L, Ciuffreda M, Maffulli N and Denaro V. Sudden cardiac death in young athletes with long QT syndrome: the role of genetic testing and cardiovascular screening. Br Med Bull. 2018;127:43–53.

5. Arunachalam K, Lakshmanan S, Maan A, Kumar N and Dominic P. Impact of Drug Induced Long QT Syndrome: A Systematic Review. J Clin Med Res. 2018;10:384–390.

6. Marstrand P, Theilade J, Andersson C, Bundgaard H, Weeke PE, Tfelt-Hansen J, Jespersen C, Gislason G, Torp-Pedersen C, Kanters JK and Jørgensen ME. Long QT syndrome is associated with an increased burden of diabetes, psychiatric and neurological comorbidities: a nationwide cohort study. Open Heart. 2019;6:e001161.

7. Yang Y, Yang Y, Liang B, Liu J, Li J, Grunnet M, Olesen SP, Rasmussen HB, Ellinor PT, Gao L, Lin X, Li L, Wang L, Xiao J, Liu Y, Liu Y, Zhang S, Liang D, Peng L, Jespersen T and Chen YH. Identification of a Kir3.4 mutation in congenital long QT syndrome. Am J Hum Genet. 2010;86:872–80.

8. Tse G, Lee S, Zhou J, Liu T, Wong ICK, Mak C, Mok NS, Jeevaratnam K, Zhang Q, Cheng SH and Wong WT. Territory-Wide Chinese Cohort of Long QT Syndrome: Random Survival Forest and Cox Analyses. Front Cardiovasc Med. 2021;8:608592.

9. Saprungruang A, Khongphatthanayothin A, Mauleekoonphairoj J, Wandee P, Kanjanauthai S, Bhuiyan ZA, Wilde AAM and Poovorawan Y. Genotype and clinical characteristics of congenital long QT syndrome in Thailand. Indian Pacing Electrophysiol J. 2018;18:165–171.

10. Gonzalez FM, Veneziano MA, Puggina A and Boccia S. A Systematic Review on the Cost-Effectiveness of Genetic and Electrocardiogram Testing for Long QT Syndrome in Infants and Young Adults. Value Health. 2015;18:700–8.

11. Perez MV, Kumarasamy NA, Owens DK, Wang PJ and Hlatky MA. Cost-effectiveness of genetic testing in family members of patients with long-QT syndrome. Circ Cardiovasc Qual Outcomes. 2011;4:76–84.

12. Quaglini S, Rognoni C, Spazzolini C, Priori SG, Mannarino S and Schwartz PJ. Cost-effectiveness of neonatal ECG screening for the long QT syndrome. Eur Heart J. 2006;27:1824–32.

13. Lee S, Zhou J, Jeevaratnam K, Wong WT, Wong ICK, Mak C, Mok NS, Liu T, Zhang Q and Tse G. Paediatric/young versus adult patients with congenital long QT syndrome or catecholaminergic polymorphic ventricular tachycardia. European Heart Journal. 2021;42:ehab724.1870.

14. Chung CT, Lee S, Zhou J, Chou OHI, Lee TTL, Leung KSK, Jeevaratnam K, Wong WT, Liu T and Tse G. Clinical Characteristics, Genetic Basis and Healthcare Resource Utilisation and Costs in Patients with Catecholaminergic Polymorphic Ventricular Tachycardia: A Retrospective Cohort Study. RCM. 2022;23.

15. Lakhani I, Zhou J, Lee S, Li KHC, Leung KSK, Hui JMH, Lee YHA, Li G, Liu T, Wong WT, Wong ICK, Mok NS, Mak CM, Zhang Q and Tse G. A Territory-Wide Study of Arrhythmogenic Right Ventricular Cardiomyopathy Patients from Hong Kong. RCM. 2022;23.

16. Lee S, Zhou J, Wong WT, Liu T, Wu WKK, Wong ICK, Zhang Q and Tse G. Glycemic and lipid variability for predicting complications and mortality in diabetes mellitus using machine learning. BMC Endocrine Disorders. 2021;21:94.

17. Lee S, Zhou J, Guo CL, Wong WT, Liu T, Wong ICK, Jeevaratnam K, Zhang Q and Tse G. Predictive scores for identifying patients with type 2 diabetes mellitus at risk of acute myocardial infarction and sudden cardiac death. Endocrinol Diabetes Metab. 2021;4:e00240.

18. Li CK, Xu Z, Ho J, Lakhani I, Liu YZ, Bazoukis G, Liu T, Wong WT, Cheng SH, Chan MT, Zhang L, Gin T, Wong MC, Wong ICK, Wu WKK, Zhang Q and Tse G. Association of NPAC score with survival after acute myocardial infarction. Atherosclerosis. 2020;301:30–36.

19. Lee S, Zhou J, Jeevaratnam K, Wong WT, Wong ICK, Mak C, Mok NS, Liu T, Zhang Q and Tse G. Paediatric/young versus adult patients with long QT syndrome. Open Heart. 2021;8.

20. Coyle K, Coyle D, Nault I, Parkash R, Healey JS, Gray CJ, Gardner MJ, Sterns LD, Essebag V, Hruczkowski T, Blier L, Wells GA, Tang ASL, Stevenson WG and Sapp JL. Cost Effectiveness of Ventricular Tachycardia Ablation Versus Escalation of Antiarrhythmic Drug Therapy: The VANISH Trial. JACC Clin Electrophysiol. 2018;4:660–668.

21. Weiss JP, Saynina O, McDonald KM, McClellan MB and Hlatky MA. Effectiveness and cost-effectiveness of implantable cardioverter defibrillators in the treatment of ventricular arrhythmias among medicare beneficiaries. Am J Med. 2002;112:519–27.

22. Mazzanti A, Trancuccio A, Kukavica D, Pagan E, Wang M, Mohsin M, Peterson D, Bagnardi V, Zareba W and Priori SG. Independent validation and clinical implications of the risk prediction model for long QT syndrome (1-2-3-LQTS-Risk). Europace. 2022;24:614–619.

23. Cho Y. Management of Patients with Long QT Syndrome. Korean Circ J. 2016;46:747–752.

24. Mönnig G, Köbe J, Löher A, Wasmer K, Milberg P, Zellerhoff S, Pott C, Zumhagen S, Radu R, Scheld HH, Haverkamp W, Schulze-Bahr E and Eckardt L. Role of implantable cardioverter defibrillator therapy in patients with acquired long QT syndrome: a long-term follow-up. Europace. 2012;14:396–401.

25. Phillips KA, Ackerman MJ, Sakowski J and Berul CI. Cost-effectiveness analysis of genetic testing for familial long QT syndrome in symptomatic index cases. Heart Rhythm. 2005;2:1294–300.

26. Manini AF, Stimmel B and Vlahov D. Racial susceptibility for QT prolongation in acute drug overdoses. J Electrocardiol. 2014;47:244–50.

